# Characteristics of patients presenting, and not presenting, to the emergency department during the early days of COVID-19

**DOI:** 10.1101/2020.05.05.20090795

**Authors:** Bjorn C. Westgard, Matthew W. Morgan, Gabriela Vazquez-Benitez, Lauren O. Erickson, Michael D. Zwank

## Abstract

**Objective:** Societal responses to the COVID-19 pandemic have had a substantial effect upon the number of patients seeking healthcare. An initial step in estimating the impact of these changes is characterizing the patients, visits, and diagnoses for whom care is being delayed or deferred.

**Methods:** We conducted an observational study, examining demographics and diagnoses for all patient visits to the ED of an urban Level-1 trauma center before and after the state declaration and compared them to visits from a similar period in 2019. We estimated the ratios of the before and after periods using Poisson regression, calculated the percent change with respect to 2019 for total ED visits, patient characteristics, and diagnoses, and then evaluated the interactions between each factor and the overall change in ED visits.

**Results:** There was a significant 35.2% drop in overall ED visits after the state declaration. Disproportionate declines were seen in visits by pediatric and older patients, women, and Medicare recipients as well as for presentations of syncope, cerebrovascular accidents, urolithiasis, abdominal and back pain. Significantly disproportionate increases were seen in ED visits for potential symptoms of COVID-19, including URIs, shortness of breath, and chest pain.

**Conclusions:** Patient concerns about health care settings and public health have significantly altered care-seeking during the COVID-19 pandemic. Overall and differential declines in ED visits for certain demographic groups and disease processes should prompt efforts to encourage care-seeking and research to monitor for the morbidity and mortality that is likely to result from delayed or deferred care.

## Introduction

In response to the spread of the novel coronavirus 2019 (COVID-19) pandemic, state governments and health systems have enacted a range of mitigation strategies and operational changes to anticipate and address an increasing number of patients with severe acute respiratory syndrome coronavirus 2 (SARS-CoV-2). At the same time, during the early days of the pandemic, health systems have also seen a decrease in the number of patients presenting for acute care unrelated to COVID-19^1^. Given public health recommendations to mitigate the spread of COVID-19, including social distancing and stay-at-home orders, many patients are not seeking and receiving care for acute conditions which may put them at significant risk for preventable morbidity and mortality in the future. The characteristics of those patients who are and are not presenting to emergency departments as a result of stay at home orders during the early days of the SARS-CoV-2 pandemic have not yet been substantially examined in the medical literature. We report changes in the characteristics of patients and presentations to the emergency department (ED) of an urban Level-1 Trauma Center before and after the statewide announcement of “peacetime emergency” public health measures to respond to the pandemic on March 13, 2020^2^.

## Methods

We conducted an observational study of visits to the ED from February 15 to April 10, 2020, a period of 28 days before through 28 days after the state’s announcement. Accounting for days of the week, we then examined a cross-section of visits from a similar period in 2019 for historical comparison. We estimated the ratios of the before and after periods using Poisson regression and calculated the percent change with respect to 2019 for the total number of ED visits and for each of the patient characteristics and diagnoses. We calculated type III p-values to evaluate the interactions between each of those factors and the overall change in ED visits. Data were collected as part of institutional operations and quality improvement and were therefore deemed by the institutional review board to be exempt from review.

## Results

After the state declaration, the ED experienced a significant decline from an average of 250 visits daily visits for the 28 days before to an average of 167 daily visits for the 28 days after (p<0.001). This represents a decline of 35.2% in ED visits overall, or a daily decline of 4 visits, with respect to 2019. When we looked at billing, there was a 23% reduction in ED charges (95% CI 19-27%) and a 13% reduction in related hospital charges (9-17%) with respect to 2019 when adjusted for inflation.

We found significant changes in the distribution of patient demographics and visit characteristics after the state declaration, particularly in patient age, gender, race, insurance, arrival mode, and disposition. There were notable decreases in ED visits by patients under age 18 (−60.1%, 95% CI −68.0 to −50.2%, p<0.001) and over age 65 (−41.3%, −33.7 to −48.0%, p<0.001), women (−40.2%, −44.4 to −35.8%, p<0.001), African American and white patients (−37.0%, −42.6 to −30.7%; −37.0%, −42.6 to −30.7%, p<0.001), Medicare recipients (−40.8%, −47.8 to −32.7%, p<0.001) and those with other insurances (e.g. liability, no-fault, workman’s compensation, −74.1%, −80.0 to −66.5%, p<0.001), ambulatory patients (−38.1%, −41.8 to −34.3%, p=0.021), and those who left prior to evaluation or discharge (−75.6%, −82.5 to −66.0%, p=0.021).

When we examined diagnoses, we noted significantly disproportionate decreases in patients presenting with syncope (−70.5%, −53.4 to −81.3%, p<0.001), cerebrovascular accidents (−58.3%, −73.3 to −34.9% p=0.049), abdominal pain (−43.3%, −50.8 to −34.8%, p=0.045), urolithiasis (-70.0%, −84.0 to −43.6%, p=0.014), and back pain (−50.7, −61.1 to −37.4%, p=0.022). We also saw significant increases in the proportion of patients presenting with upper respiratory infections (URIs, −10.0%, −29.4 to 14.8%, p=0.007), shortness of breath (25.1%, −6.4 to 67.3%, p<0.001), and chest pain (−13.1%, −27.2 to 3.7%, p<0.001). Of note, proportional increases were also seen for all of the mental health and substance-use diagnoses we examined, though these were not significantly disproportionate. For all other conditions, the declines in presentations were proportionate to the overall change in ED visits.

## Discussion

Paralleling anecdotal reports, we noted a significant temporal association between our state’s emergency declaration and an overall decline in daily ED visits. We found significant changes in ED patient demographics. The significant proportional decreases in visits by the overlapping populations of patients over 65 and Medicare recipients are likely a product of the extra prudence of higher risk individuals who are social distancing and staying at home. The significant proportional declines in visits by pediatric, ambulatory, and other patients likely reflect concerns about contracting COVID-19 in health care settings, overburdening the health care system with unrelated complaints, and adhering to public health recommendations. The decrease in number of patients who left prior to evaluation or discharge is likely due at least in part to the substantial operational changes made in response to the pandemic which have further streamlined patient triage, rooming, and evaluation.

The most alarming finding of this report, however, is simply the overall decline in care-seeking for acute and potentially life-threatening conditions unrelated to COVID-19^1^. Societal responses to the COVID-19 pandemic have contributed to multiple phenomena, including diminished air pollution, traffic, and infectious disease transmission, which could all potentially contribute to improved population health over the long term^1^. These phenomena may well influence exacerbations of respiratory conditions as well as COVID-19 infections and outcomes over the short term^3^, but most of the conditions seen and managed in the emergency department would not be expected to suddenly decrease in incidence. Variable disease severity could play a role, as patients with less acute conditions might be able to delay or defer seeking care. Though we did not examine low acuity presentations that might also be managed in other outpatient settings, we did find a disproportionate decrease in presentations of back pain and other non-specific pains. However, we also found unexpectedly disproportionate declines in visits for conditions of significantly higher acuity like syncope, cerebrovascular accidents, and urolithiasis, similar to those noted elsewhere for presentations of myocardial infarction^4,5^. Additionally, we saw increases in ED visits for upper respiratory infections, chest pain, or shortness of breath, symptoms that might bring patients to the ED with concerns for COVID-19 infection. Similar trends in patient presentations have been seen around the world^6^, during prior disease outbreaks^8^, and in the outpatient setting^8^.

These are early days, and the timeline for the COVID-19 pandemic and resulting changes in patterns of ED utilization promise to be much longer than that covered in our preliminary study. This study is limited by its single center and cross-sectional nature, the short study time frame, and lack of adjustment for multiple potentially confounding factors related to patients and their presentations to the ED, including disease severity, comorbidities, and COVID-19 risk factors. In spite of those limitations, we have found notable differential changes in the demographic factors, visits characteristics, and diagnoses of presentations to the ED unrelated to COVID-19. These will no doubt change along with the burdens of care as the pandemic progresses. Further efforts should and are being made to reassure and affirm the appropriateness of seeking emergency care^9^, particularly for the groups and disease processes that have been highlighted here and elsewhere^4,5^. Further research will also be needed to examine these and other factors contributing to delayed or deferred care and to monitor for the morbidity and mortality that is likely to result^5,8^ and which may already be occurring^10^.

**Figure 1.**
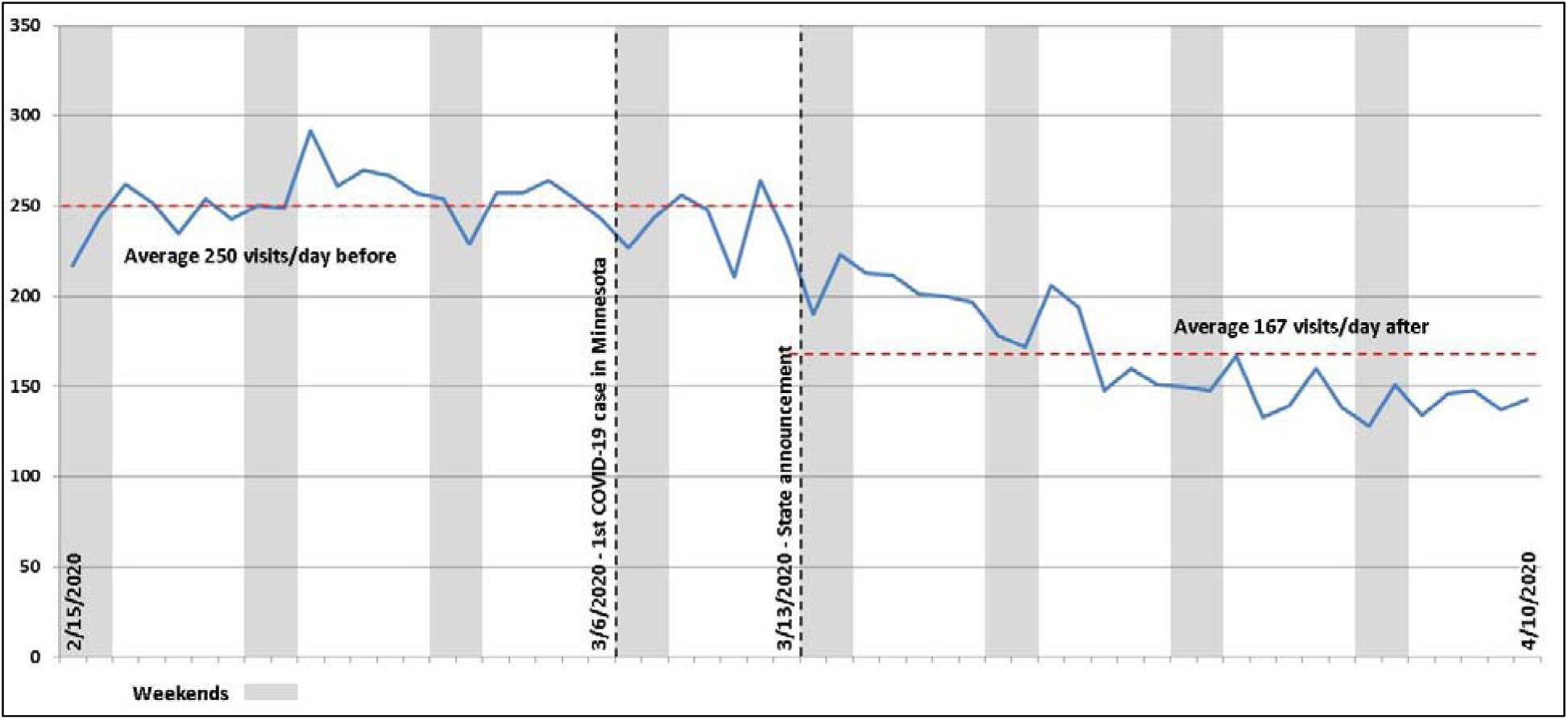
Trend of daily ED visits before and after state announcement.

**Table 1.**
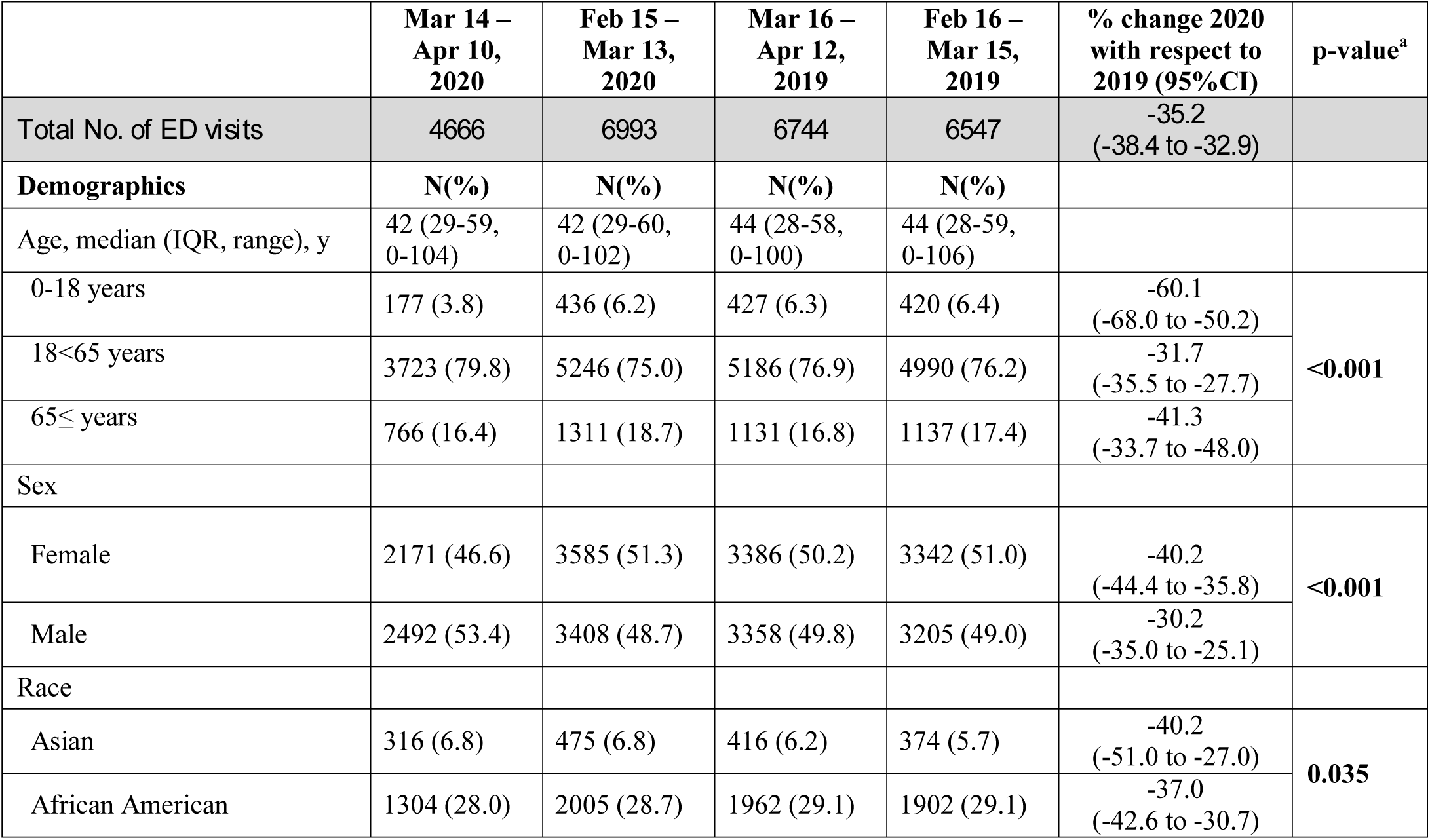
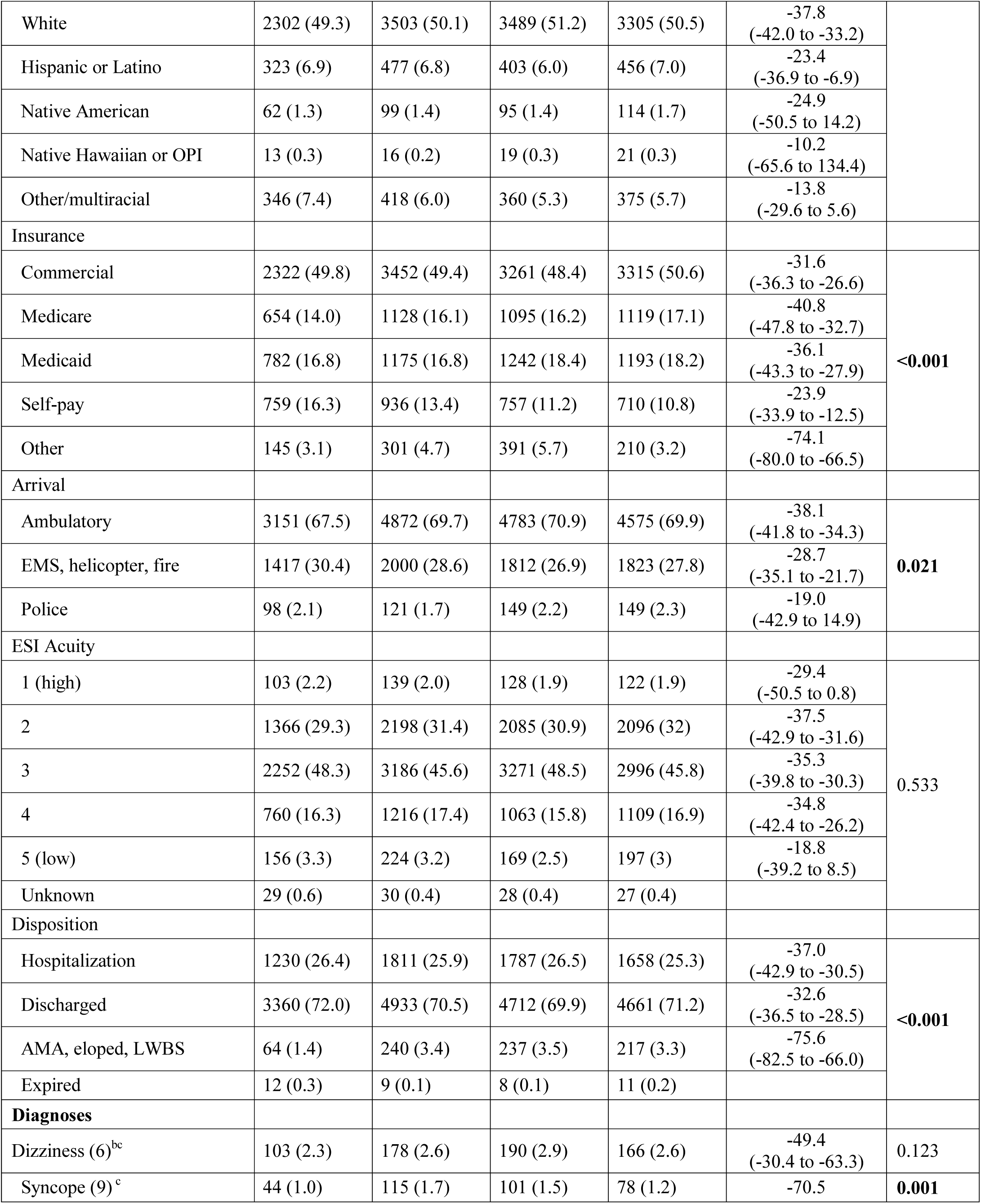
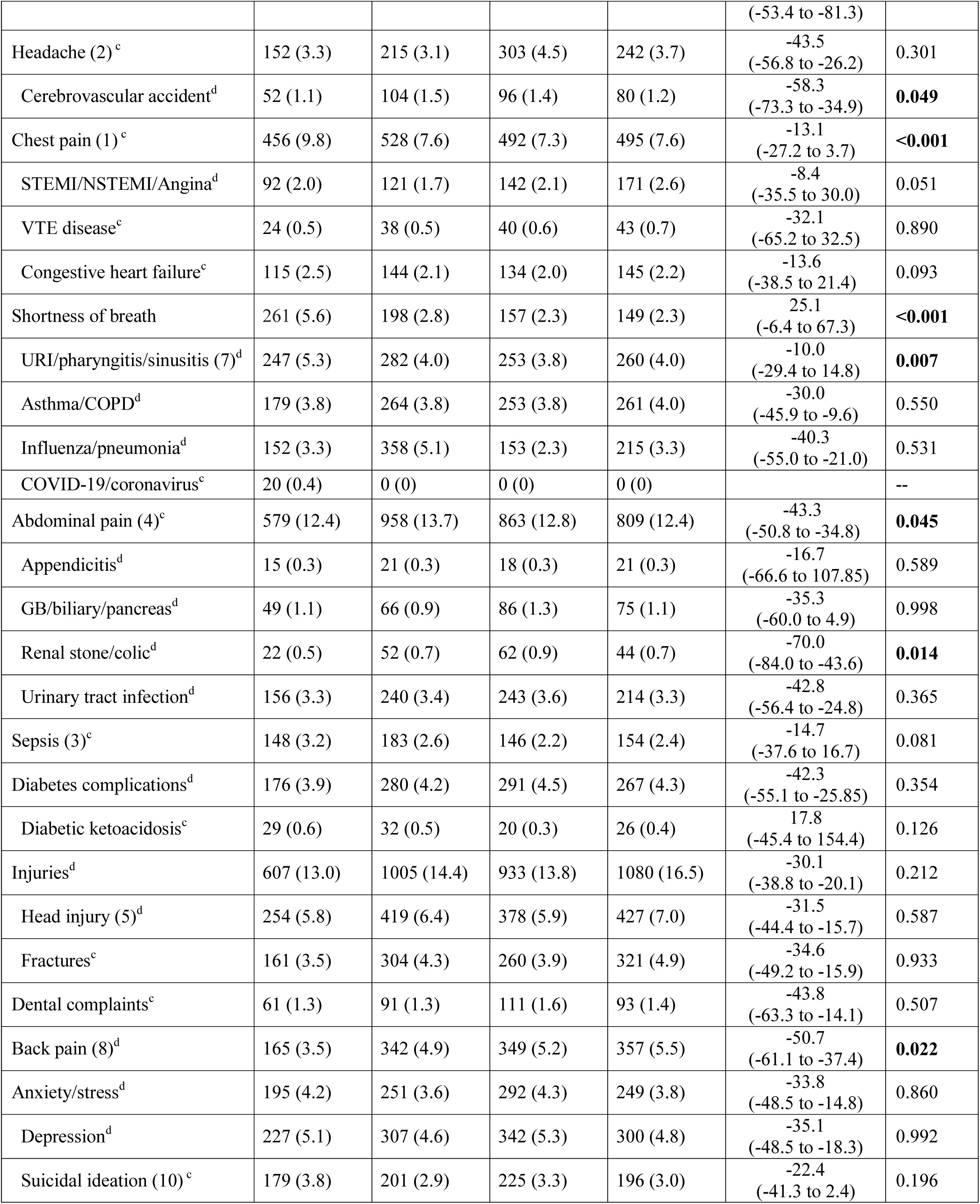
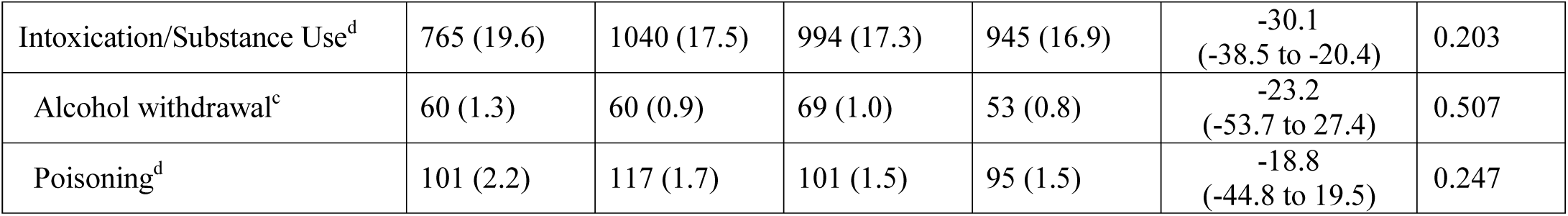
Characteristics of ED patients and presentations before and after state announcement. Abbreviations: IQR, inter-quartile range; OPI, other Pacific Islander; EMS, Emergency Medical Services; ESI, Emergency Severity Index; AMA, against medical advice; LWBS, left without being seen; STEMI, ST-elevation myocardial infarction; NSTEMI, non-ST-elevation myocardial infarction; VTE, venous thromboembolic; URI, upper respiratory infection; COPD, chronic obstructive pulmonary disease; COVID-19, novel coronavirus disease 2019; and GB, gallbladder. ^a^ P-value represents the type III p-value for the interaction between the overall change in ED visits and the change in each category of patient and visit characteristics and diagnoses. ^b^ Numbers indicate historical top 10 diagnoses for visits to our ED. ^c^ Determined from a search of diagnosis names within the first 3 coder diagnoses. ^d^ Determined from a search of diagnostic related groups within the first 3 coder diagnoses.

## Data Availability

Limited data is available if necessary, but the data as currently analyzed does include PHI.

## Acknowledgement

We would like to thank Hrafn P. Gudjonsson (“Patrick”) for his valuable assistance in querying and creating the data set used in this study.

---

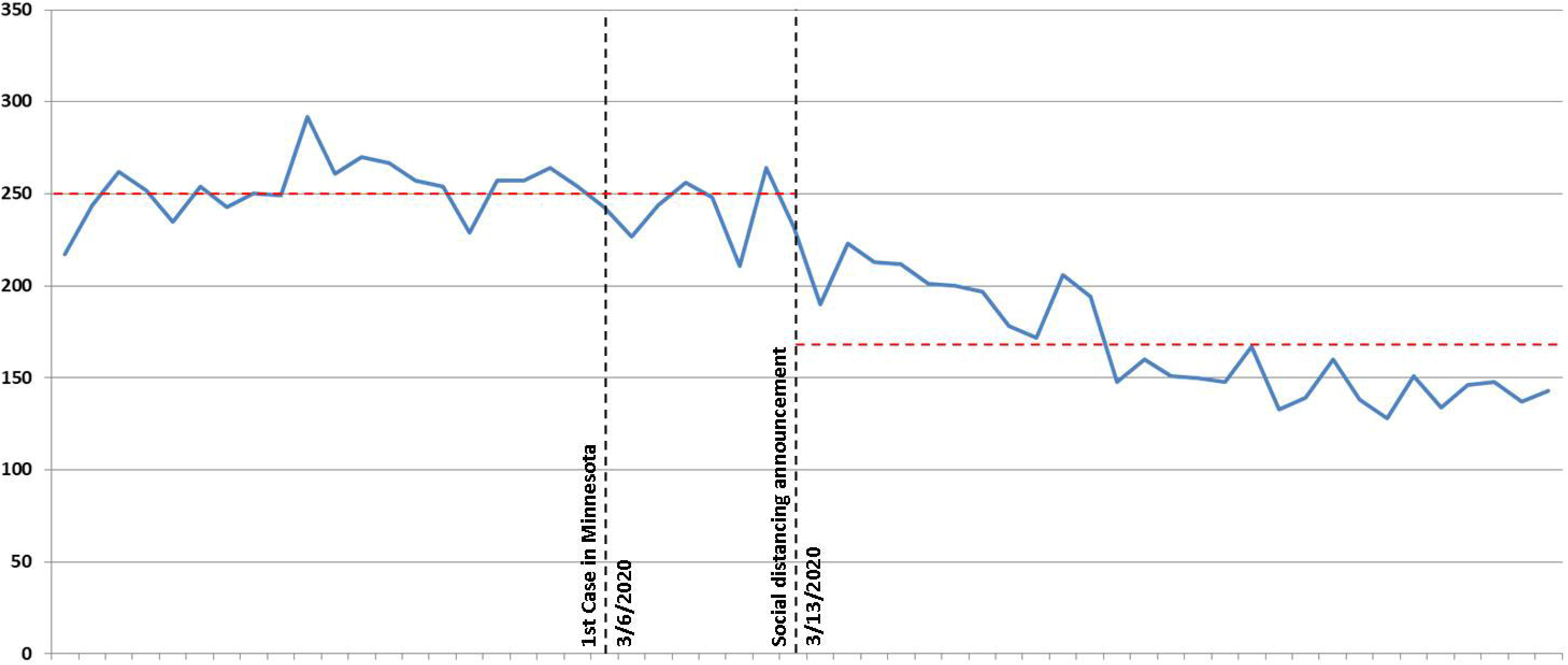

